# A cross-sectional analysis of demographic and behavioral risk factors of SARS-CoV-2 antibody positivity among a sample of U.S. college students

**DOI:** 10.1101/2021.01.20.21249905

**Authors:** Sina Kianersi, Christina Ludema, Jonathan T. Macy, Edlin Garcia, Chen Chen, Maya Luetke, Mason H. Lown, Molly Rosenberg

## Abstract

**Background:** Colleges and universities across the United States are developing and implementing data-driven prevention and containment measures against severe acute respiratory syndrome coronavirus 2 (SARS-CoV-2) infection. Identifying risk factors for SARS-CoV-2 seropositivity could help to direct these efforts.

**Objective:** To estimate the associations between demographic factors and social behaviors and SARS-CoV-2 seropositivity and self-reported positive SARS-CoV-2 diagnostic test.

**Methods:** In September 2020, we randomly sampled Indiana University Bloomington (IUB) undergraduate students. Participants completed a cross-sectional, online survey about demographics, SARS-CoV-2 testing history, relationship status, and risk behaviors. Additionally, during a subsequent appointment, participants were tested for SARS-CoV-2 antibodies using a fingerstick procedure and SARS-CoV-2 IgM/IgG rapid assay kit. We used unadjusted modified Poisson regression models to evaluate the associations between predictors of both SARS-CoV-2 seropositivity and self-reported positive SARS-CoV-2 infection history.

**Results:** Overall, 1,076 students were included in the serological testing analysis, and 1,239 students were included in the SARS-CoV-2 infection history analysis. Current seroprevalence of SARS-CoV-2 was 4.6% (95% CI: 3.3%, 5.8%). Prevalence of self-reported SARS-CoV-2 infection history was 10.3% (95% CI: 8.6%, 12.0%). Greek membership, having multiple romantic partners, knowing someone in one’s immediate environment with SARS-CoV-2 infection, drinking alcohol more than 1 day per week, and hanging out with more than 4 people when drinking alcohol increased both the likelihood of seropositivity and SARS-CoV-2 infection history.

**Conclusion:** Our findings have implications for American colleges and universities and could be used to inform SARS-C0V-2 prevention and control strategies on such campuses.

## 1 Introduction

### 1.1 Background and rationale

The ongoing Coronavirus Disease 2019 (COVID-19) pandemic has caused major challenges for both colleges and students including school closures, shifts to remote and hybrid educational formats, and negative financial impacts (1). More importantly, the disease burden on college campuses has been substantial with at least 397,000 severe acute respiratory syndrome coronavirus 2 (SARS-CoV-2) cases and at least 90 deaths reported at more than 1,900 colleges as of December 11, 2020 (2). Furthermore, because of the collegiate semester schedules, there are mounting concerns that infected asymptomatic students might spread the virus to their family members when traveling back home (3). Identifying predictors of SARS-CoV-2 test positivity can help to plan and coordinate mitigation testing programs, containment efforts, and vaccination strategies.

Previous studies among the general U.S. adult population have established that race, gender, and age are associated with SARS-CoV-2 positivity (4, 5). However, these characteristics have not been thoroughly studied among college students. Moreover, there are demographic factors specific to college student populations, such as participation in Greek life (6), dating, and year in school, that might be significant predictors of SARS-CoV-2 positivity in this population.

Lastly, because of the drinking culture and social context of drinking among college students (7), alcohol use patterns may be another potential predictor of SARS-CoV-2 positivity in this population. Young adults with a hazardous drinking problem have reported to comply less with the stay-at-home order on days that they were drinking, compared to days that they did not drink (8). The effects of alcohol are compounded in the social setting of college drinking: when the number of friends present in an alcohol drinking event increases, the number of consumed alcohol drinks increases (at an individual level) (9). Crowded social events also, by definition, limit the ability to maintain physical distance. Since alcohol consumption is prevalent among college students (10), assessing the relationship between this behavior and SARS-CoV-2 positivity is imperative to better understand the dynamics of SARS-CoV-2 transmission among college students. Therefore, in the current study we examined the relationship between drinking behaviors and SARS-CoV-2 positivity.

### 1.2 Objective

The primary aim of the current study was to estimate the associations between different demographic characteristics and social behaviors and SARS-CoV-2 seropositivity and self-reported positive test history outcomes among college students. We also estimated the seroprevalence of SARS-CoV-2 antibody (in September 2020) and the prevalence of self-reported SARS-CoV-2 positive test history among Indiana University Bloomington (IUB) undergraduate students.

## 2 Methods

We used the Strengthening the Reporting of Observational Studies in Epidemiology (STROBE) guidelines (11) to report our findings about the baseline characteristics of IUB COVID-19 Serosurvey Study participants and predictors of SARS-CoV-2 positivity.

### 2.1 Study Design

The parent study design was a randomized controlled trial (RCT) to test whether receiving SARS-CoV-2 antibody test results alters students’ protective behaviors against infection (12). The RCT data collection was longitudinal and lasted for two months. We collected data at baseline and every two weeks post-baseline. In the current study, we used data from baseline survey and baseline antibody test results in a cross-sectional study design. Participants were compensated with a maximum of $30 USD for completing all the steps in the RCT. This study protocol #2008293852 received approval from the University’s Institutional Review Board.

### 2.2 Setting

Study invitation emails were sent to a random sample of 7,499 IUB undergraduate students. The emails included information about the study and a student-specific link to an online survey. The online survey consisted of an eligibility criteria instrument, an online consent form, a laboratory test appointment scheduler for SARS-CoV-2 antibody test, and a baseline survey. The baseline survey measured participants demographics, SARS-CoV-2 testing history, and risk behaviors. Eligible students who consented to participate were able to schedule a laboratory test appointment and complete the online baseline survey. Study invitation and reminder emails were sent on September 8-20, 2020. Students scheduled their baseline appointments and responded to the baseline survey between September 8 and September 30, 2020.

The SARS-CoV-2 antibody laboratory tests were conducted in-person outdoors on the IUB campus, between September 14 - 30. During laboratory test, recommended protocols to reduce the likelihood of SARS-CoV-2 transmission at the study site were employed, including physical distancing, mask wearing, glove wearing, and disinfection of laboratory equipment. Students were advised not to attend their appointment if they were experiencing COVID-19 symptoms, had tested positive for SARS-CoV-2 in the last two weeks before their appointment, or had been directed to isolate or quarantine. Participants checked in with their unique study ID, which they had created in the online survey. Using a fingerstick procedure, trained nursing staff took a small blood sample from each participant and placed the blood sample on the antibody testing kit. Trained field staff read the antibody test results from the test kit, took a high-quality picture of the kit and uploaded it to a secured cloud drive, and entered the test results into the REDCap data management system. To increase the accuracy of the antibody test readings, a trained research assistant independently assessed the results using the pictures that field staff had taken from the test kits. Discordant results were adjudicated by five research team members.

### 2.3 Participants

We selected a random sample of IUB undergraduate students (n=7,499) from the sampling frame of all IUB undergraduate students (n∼33,084). Selected students were eligible to participate in this study if they were 1) age 18 or older, 2) a current IUB undergraduate student, and 3) currently residing in Monroe County, IN.

### 2.4 Variables and data sources/measurement

#### 2.4.1 Outcomes

Objective outcome: The main outcome was the participants’ SARS-CoV-2 antibody laboratory test result. The virus can cause immune response in both symptomatic and asymptomatic individuals (13, 14). The antibody test kits we used were SARS-CoV-2 IgM/IgG rapid assay kit (Colloidal Gold method). These kits can detect IgM and IgG antibodies against SARS-CoV-2 in the blood and provide accurate and rapid results at the testing site. If the antibody test result was negative for both IgM and IgG antibodies, the antibody test result was coded as negative.

Otherwise, if the test kit results for any of the two types of antibodies was positive, the outcome was coded positive.

Subjective outcome: The second outcome of interest was self-reported SARS-CoV-2 testing history. This was measured by the following questions in the baseline survey.

1. “*Have you ever been tested for SARS-CoV-2 (COVID-19) before? Note: By this, we mean testing for active infections, usually done with a nasal swab or saliva test”* (Responses: “Yes”, “No”, “Don’t Know”).
2. [Displayed if 1 equals Yes] “*Have you ever tested positive for a SARS-CoV-2 (COVID-19) infection?”* (Responses: “Yes”, “No”, “Don’t Know”)

Participants who responded “Yes” to both questions were categorized as ever tested positive for SARS-CoV-2.

#### 2.4.2 Demographic and behavioral predictors

We collected data on the following baseline characteristics and potential risk factors for a positive SARS-CoV-2 test result: age (≥22 years old vs. <22 years old), sex at birth (Female vs. Male), race (Asian, Black, Multi-racial, Other, White), Hispanic or Latinx ethnicity (Yes vs. No), year in school (1^st^ through 5^th^), residence (On-campus vs. Off-campus), Greek membership (Yes vs. No), relationship status (Multiple partners, Single partner, No partner), know others who were infected (Yes vs. No), and number of days per week drinking alcohol (>1 day in a week vs. ≤1 day in a week). Moreover, among those who self-reported a positive SARS-CoV-2 testing history we collected data about their symptoms, symptomatic (Yes vs. No), and among students who reported drinking alcohol, we collected data about the number of people they hung out with while drinking (>4 people vs. ≤4 people). All the above-mentioned predictors were self-reported in the online baseline survey (Appendix A).

Three of the above-mentioned variables were collected as continuous variables but were dichotomized for analyses: (1) Age: Undergraduate students are mainly 18-22 years old. We recoded this continuous variable as categorical with 22 years old as the cut-off point. (2) Number of days per week drinking alcohol: This continuous variable could range from 0 to 7. We used the median of one as the cut-off point for this variable. (3) Number of people hanging out with while drinking: This continuous variable could range from 0 to 1,000. We used the median of four as the cut-off point.

### 2.5 Bias

We took several measures to reduce different sources of bias, such as non-response and selection biases. We used a random sample to decrease selection bias. Besides the initial study invitation email, we sent two reminders to participants to increase the response rate. We also identified different types of partial responses and sent reminder emails to participants who had only completed part of the baseline study. Moreover, to maximize the number of participants showing up for their SARS-CoV-2 antibody testing appointment, we sent appointment reminders to participants 6-12 hours before their appointments.

### 2.6 Study Size

The sample size calculation for the RCT was calculated before conducting that study. However, the power analysis was specific to the RCT aims, and therefore no sample size calculation was conducted for the current cross-sectional analysis of the baseline data.

### 2.7 Statistical Methods

We used the normal approximation interval (Wald interval) to calculate the point prevalence/seroprevalence estimates and 95% confidence intervals (CI) for positive SARS-CoV-2 antibody test and self-reported history of positive SARS-CoV-2 test. We used Poisson regression models with a robust error variance to calculate the unadjusted/crude associations between different baseline variables and the self-reported SARS-CoV-2 testing history and SARS-CoV-2 antibody laboratory test outcome variables (15, 16). We report the unadjusted prevalence ratios and 95% CIs for these associations. Values of “Don’t Know” were recoded as missing in the analysis. We used complete case analysis. Lastly, in a sensitivity analysis, to remove any biases that age outliers were potentially introducing to our findings, we restricted our sample to students below 30 years old and ran the models.

## 3 Results

### 3.1 Participants

We sampled 7,499 IUB undergraduate students, 4,069 students were likely eligible based on county of residence, 1,397 confirmed eligibility and consented to participate in the study, and 1,076 attended a laboratory test visit and provided SARS-CoV-2 antibody test data. Overall, 21 students explicitly refused to participate in the study while 2,651 tacitly refused via non-response or by not signing the consent form. Moreover, among students who consented to participate in the study, 321 students did not schedule or attend a baseline antibody test appointment.

For the self-reported SARS-CoV-2 testing history outcome, out of the 1,397 students who consented to participate in the study, 133 did not answer any of the questions in the baseline survey, and 25 students had missing values for the self-reported outcome. Overall, 1,239 answered the survey questions about SARS-CoV-2 testing history (Figure 1). We calculated the response rate to be 26.4% for the antibody testing outcome and 30.4% for the self-reported SARS-CoV-2 testing history outcome.

**Figure 1.**
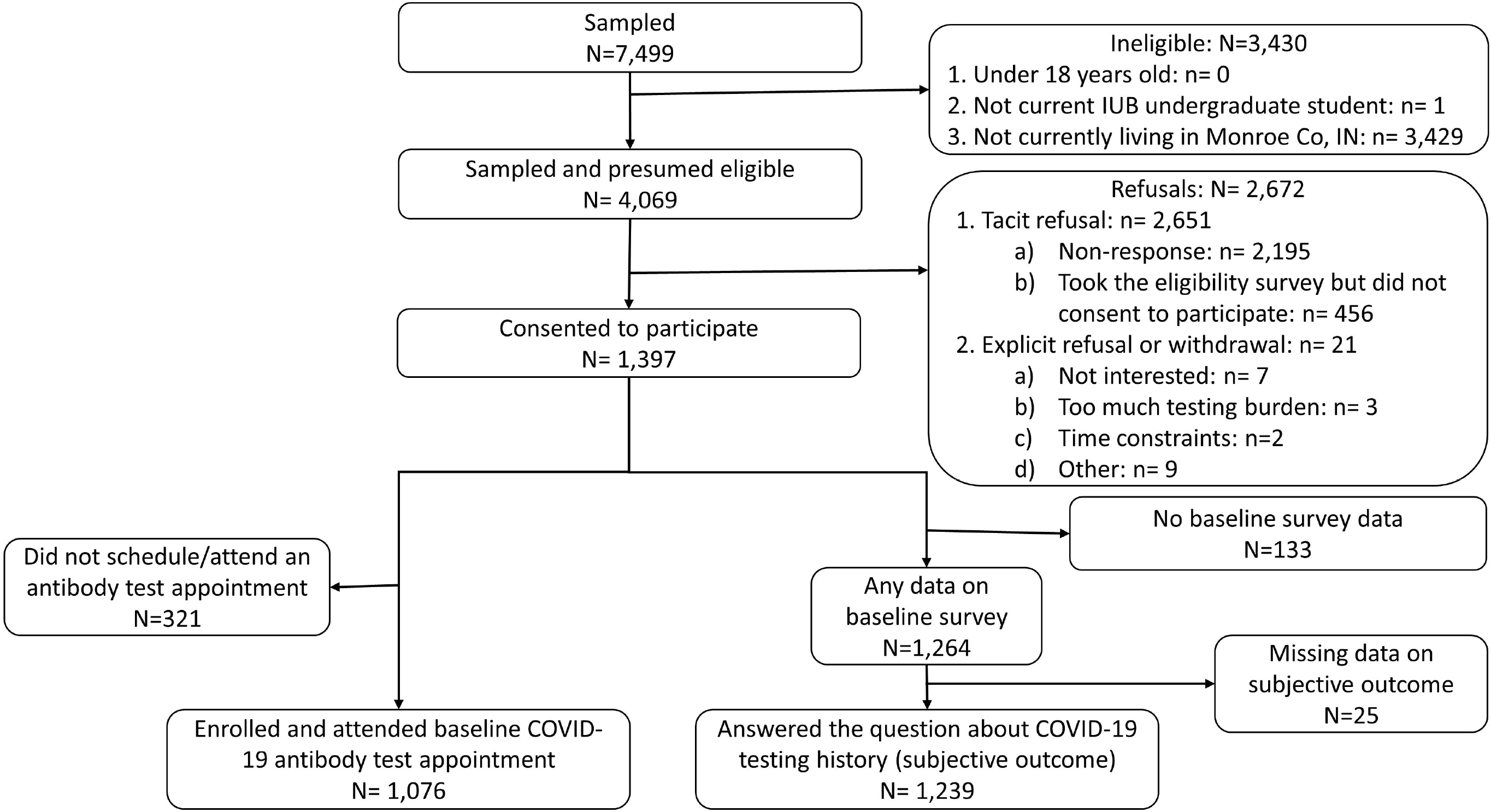
Flow diagram of the study sample

### 3.2 Descriptive Data

For the SARS-CoV-2 antibody laboratory test outcome, participants were on average 20 years old (Median, Standard Deviation: 20, 2.5), female (64%), white (79%), non-Hispanic (93%), senior student (28%), off-campus residents (69%), non-Greek affiliated (76%), and single and not dating/hooking up with anyone (40%). Moreover, 49% of participants knew others with SARS-CoV-2 positive history, 46% reported drinking alcohol >1 day per week, and 41% reported hanging out with >4 people while drinking (Table 1). Similar trends in demographic and behavioral variables were found for the self-reported SARS-CoV-2 testing history outcome.

**Table 1.** Characteristics of the study participants, Indiana University Bloomington undergraduate students, September 2020

|  | Antibody test |  |  |  |  |  | Self-reported positive SARS-CoV-2 test |  |  |  |  |  |
| --- | --- | --- | --- | --- | --- | --- | --- | --- | --- | --- | --- | --- |
|  | Overall<br>1076 |  | Negative<br>1027 |  | Positive<br>49 |  | Overall<br>1239 |  | No<br>1111 |  | Yes<br>128 |  |
|  | N | (%) | N | (%) | N | (%) | N | (%) | N | (%) | N | (%) |
| **Age** |  |  |  |  |  |  |  |  |  |  |  |  |
| 18 years old | 208 | (20.6) | 198 | (20.5) | 10 | (21.7) | 247 | (21.2) | 217 | (20.9) | 30 | (24.0) |
| 19 years old | 224 | (22.2) | 215 | (22.3) | 9 | (19.6) | 253 | (21.7) | 223 | (21.5) | 30 | (24.0) |
| 20 years old | 228 | (22.6) | 219 | (22.7) | 9 | (19.6) | 254 | (21.8) | 231 | (22.2) | 23 | (18.4) |
| 21 years old | 255 | (25.3) | 241 | (25.0) | 14 | (30.4) | 297 | (25.5) | 263 | (25.3) | 34 | (27.2) |
| 22+ years old | 95 | (9.4) | 91 | (9.4) | 4 | (8.7) | 113 | (9.7) | 105 | (10.1) | 8 | (6.4) |
| Missing | 66 |  | 63 |  | 3 |  | 75 |  | 72 |  | 3 |  |
| **Sex at birth** |  |  |  |  |  |  |  |  |  |  |  |  |
| Female | 689 | (64.3) | 655 | (64.0) | 34 | (70.8) | 786 | (63.5) | 709 | (63.8) | 77 | (60.6) |
| Male | 382 | (35.7) | 368 | (36.0) | 14 | (29.2) | 452 | (36.5) | 402 | (36.2) | 50 | (39.4) |
| Missing | 5 |  | 4 |  | 1 |  | 1 |  | 0 |  | 1 |  |
| **Race** |  |  |  |  |  |  |  |  |  |  |  |  |
| Asian | 80 | (7.5) | 77 | (7.5) | 3 | (6.1) | 95 | (7.7) | 90 | (8.1) | 5 | (3.9) |
| Black | 13 | (1.2) | 13 | (1.3) | 0 | (0.0) | 24 | (1.9) | 24 | (2.2) | 0 | (0.0) |
| Multi-racial | 85 | (7.9) | 80 | (7.8) | 5 | (10.2) | 101 | (8.2) | 84 | (7.6) | 17 | (13.3) |
| Other | 46 | (4.3) | 43 | (4.2) | 3 | (6.1) | 59 | (4.8) | 58 | (5.2) | 1 | (0.8) |
| White | 847 | (79.1) | 809 | (79.2) | 38 | (77.6) | 959 | (77.5) | 854 | (76.9) | 105 | (82.0) |
| Missing | 5 |  | 5 |  | 0 |  | 1 |  | 1 |  | 0 |  |
| **Hispanic or Latinx Ethnicity** |  |  |  |  |  |  |  |  |  |  |  |  |
| No | 998 | (92.8) | 955 | (93.0) | 43 | (87.8) | 1146 | (92.5) | 1028 | (92.5) | 118 | (92.2) |
| Yes | 78 | (7.3) | 72 | (7.0) | 6 | (12.2) | 93 | (7.5) | 83 | (7.5) | 10 | (7.8) |
| **Year in school** |  |  |  |  |  |  |  |  |  |  |  |  |
| 1st | 236 | (22.1) | 224 | (21.9) | 12 | (24.5) | 280 | (22.7) | 249 | (22.5) | 31 | (24.4) |
| 2nd | 246 | (23.0) | 235 | (23.0) | 11 | (22.4) | 277 | (22.4) | 245 | (22.1) | 32 | (25.2) |
| 3rd | 264 | (24.7) | 255 | (25.0) | 9 | (18.4) | 302 | (24.4) | 276 | (24.9) | 26 | (20.5) |
| 4th | 297 | (27.8) | 281 | (27.5) | 16 | (32.7) | 343 | (27.8) | 306 | (27.6) | 37 | (29.1) |
| 5th | 27 | (2.5) | 26 | (2.5) | 1 | (2.0) | 34 | (2.8) | 33 | (3.0) | 1 | (0.8) |
| Missing | 6 |  | 6 |  | 0 | 0 | 3 |  | 2 |  | 1 |  |
| **Residence** |  |  |  |  |  |  |  |  |  |  |  |  |
| Off-campus | 738 | (68.9) | 705 | (69.0) | 33 | (67.3) | 844 | (68.2) | 764 | (68.8) | 80 | (62.5) |
| On-campus | 333 | (31.1) | 317 | (31.0) | 16 | (32.7) | 394 | (31.8) | 346 | (31.2) | 48 | (37.5) |
| Missing | 5 |  | 5 |  | 0 |  | 1 |  | 1 |  | 0 |  |
| **Greek membership** |  |  |  |  |  |  |  |  |  |  |  |  |
| No | 812 | (75.9) | 788 | (77.2) | 24 | (49.0) | 943 | (76.3) | 870 | (78.5) | 73 | (57.0) |
| Yes | 258 | (24.1) | 233 | (22.8) | 25 | (51.0) | 293 | (23.7) | 238 | (21.5) | 55 | (43.0) |
| Missing | 6 |  | 6 |  | 0 |  | 3 |  | 3 |  | 0 |  |
| **Relationship status** |  |  |  |  |  |  |  |  |  |  |  |  |
| Single and not dating/hooking up with anyone | 432 | (40.3) | 415 | (40.6) | 17 | (34.7) | 499 | (40.3) | 448 | (40.4) | 51 | (39.8) |
| Single and dating/hooking up with multiple people | 101 | (9.4) | 91 | (8.9) | 10 | (20.4) | 121 | (9.8) | 94 | (8.5) | 27 | (21.1) |
| Single and dating/hooking up with one specific person | 138 | (12.9) | 133 | (13.0) | 5 | (10.2) | 165 | (13.3) | 151 | (13.6) | 14 | (10.9) |
| In a relationship but not living together | 358 | (33.4) | 342 | (33.4) | 16 | (32.7) | 404 | (32.6) | 368 | (33.2) | 36 | (28.1) |
| Living together but not married | 40 | (3.7) | 40 | (3.9) | 0 | (0.0) | 46 | (3.7) | 46 | (4.1) | 0 | (0.0) |
| Married and living together | 3 | (0.3) | 2 | (0.2) | 1 | (2.0) | 3 | (0.2) | 3 | (0.3) | 0 | (0.0) |
| Missing | 4 |  | 4 |  | 0 |  | 1 |  | 1 |  | 0 |  |
| **Self-report positive test** |  |  |  |  |  |  |  |  |  |  |  |  |
| No | 961 | (90.8) | 944 | (93.2) | 17 | (37.0) | -- | -- | -- | -- | -- | -- |
| Yes | 98 | (9.3) | 69 | (6.8) | 29 | (63.0) | -- | -- | -- | -- | -- | -- |
| Missing | 17 |  | 14 |  | 3 |  | -- | -- | -- | -- | -- | -- |
| **Symptomatic <sup>a</sup>** |  |  |  |  |  |  |  |  |  |  |  |  |
| No | 23 | (24.0) | 18 | (26.9) | 5 | (17.2) | 31 | (24.6) | 0 | (0.0) | 31 | (24.6) |
| Yes | 73 | (76.0) | 49 | (73.1) | 24 | (82.8) | 95 | (75.4) | 0 | (0.0) | 95 | (75.4) |
| Missing | 980 |  | 960 |  | 20 |  | 1113 |  | 1111 |  | 2 |  |
| **Know others who were infected** |  |  |  |  |  |  |  |  |  |  |  |  |
| No | 510 | (49.4) | 501 | (50.9) | 9 | (18.8) | 581 | (48.6) | 551 | (51.5) | 30 | (23.8) |
| Yes | 523 | (50.6) | 484 | (49.1) | 39 | (81.3) | 614 | (51.4) | 518 | (48.5) | 96 | (76.2) |
| Missing | 43 |  | 42 |  | 1 |  | 44 |  | 42 |  | 2 |  |
| **Number of days in a week drinking alcohol** |  |  |  |  |  |  |  |  |  |  |  |  |
| 1 day or less | 577 | (54.0) | 558 | (54.7) | 19 | (39.6) | 652 | (53.3) | 600 | (54.6) | 52 | (41.6) |
| More than 1 day | 491 | (46.0) | 462 | (45.3) | 29 | (60.4) | 572 | (46.7) | 499 | (45.4) | 73 | (58.4) |
| Missing | 8 |  | 7 |  | 1 |  | 15 |  | 12 |  | 3 |  |
| **Number of people hanging out with while drinking** |  |  |  |  |  |  |  |  |  |  |  |  |
| 4 people or less | 615 | (59.2) | 596 | (60.1) | 19 | (39.6) | 702 | (59.0) | 648 | (60.7) | 54 | (44.6) |
| More than 4 people | 424 | (40.8) | 395 | (39.9) | 29 | (60.4) | 487 | (41.0) | 420 | (39.3) | 67 | (55.4) |
| Missing | 37 |  | 36 |  | 1 |  | 50 |  | 43 |  | 7 |  |
| a. Among those who self-reported a positive test |  |  |  |  |  |  |  |  |  |  |  |  |

### 3.3 Outcome Data

Overall, 49 students (out of 1076) tested positive for SARS-CoV-2 antibodies [Prevalence (95% CI): 4.6% (3.3%, 5.8%)] and 128 students (out of 1239) self-reported ever having tested positive for SARS-CoV-2 infection [Prevalence (95% CI): 10.3% (8.6%, 12.0%)] (Table 2).

**Table 2.** Bivariate prevalence ratios for the associations between risk factors and positive SARS-CoV-2 antibody test and self-reported history of positive SARS-CoV-2 test.

| Predictor | Outcomes |  |
| --- | --- | --- |
|  | Positive SARS-CoV-2 Antibody Test (objective outcome) | Self-reported History of Positive SARS-CoV-2 Test (subjective outcome) |
|  | PR (95% CI) | PR (95% CI) |
| **Age** |  |  |
| ≥22 years old | 0.92 (0.34, 2.50) | 0.63 (0.32, 1.26) |
| <22 years old | Ref. | Ref. |
| **Sex at birth** |  |  |
| Female | 1.35 (0.73, 2.48) | 0.89 (0.63, 1.24) |
| Male | Ref. | Ref. |
| **Hispanic or Latinx Ethnicity** |  |  |
| Yes | 1.79 (0.78, 4.06) | 1.04 (0.57, 1.92) |
| No | Ref. | Ref. |
| **Year in school** |  |  |
| 1<sup>st</sup> | 0.97 (0.47, 1.99) | 1.10 (0.70, 1.72) |
| 2<sup>nd</sup> | 0.85 (0.41, 1.79) | 1.15 (0.74, 1.79) |
| 3<sup>rd</sup> | 0.65 (0.29, 1.43) | 0.85 (0.53, 1.37) |
| 4<sup>th</sup> or 5<sup>th</sup> | Ref. | Ref. |
| **Residence** |  |  |
| On -campus | 1.07 (0.60, 1.92) | 1.29 (0.92, 1.80) |
| Off-campus | Ref. | Ref. |
| **Greek membership** |  |  |
| Yes | **3.28 (1.91, 5.64)** | **2.42 (1.75, 3.35)** |
| No | Ref. | Ref. |
| **Relationship Status** |  |  |
| Multiple partners | **2.52 (1.19, 5.33)** | **2.18 (1.43, 3.33)** |
| Single partner | 1.04 (0.5, 1.93) | 0.79 (0.55, 1.15) |
| No partner | Ref. | Ref. |
| **Self-report positive test (Subjective outcome)** |  |  |
| Yes | **16.73 (9.54, 29.33)** | -- |
| No | Ref. | -- |
| **Symptomatic <sup>a</sup>** |  |  |
| Yes | 1.51 (0.65, 3.51) | -- |
| No | Ref. | -- |
| **Know others who were infected** |  |  |
| Yes | **4.23 (2.07, 8.63)** | **3.03 (2.04, 4.49)** |
| No | Ref. | Ref. |
| **Number of days per week drinking alcohol** |  |  |
| More than 1 day | **1.79 (1.02, 3.16)** | **1.60 (1.14, 2.24)** |
| 1 day or less | Ref. | Ref. |
| **Number of people hanging out with while drinking** |  |  |
| More than 4 people | **2.21 (1.26, 3.90)** | **1.79 (1.27, 2.51)** |
| 4 people or less | Ref. | Ref. |
| a. Displayed if self-report positive test = Yes |  |  |
| Boldface indicates $p < 0.05$ | | |

### 3.4 Main Results

#### 3.4.1 Objective outcome: SARS-CoV-2 antibody laboratory test

Students affiliated with Greek fraternities or sororities were 3.28 (95% CI: 1.91, 5.64) times more likely to have a positive SARS-CoV-2 antibody test result compared to non-Greek students. Those with multiple partners were 2.52 (95% CI: 1.19, 5.33) times more likely to have a positive antibody test compared to students with no partner. However, those with single partners had similar distribution of positive antibody tests compared to those with no partner [PR (95% CI): 1.04 (0.5, 1.93)]. Students who knew others in their immediate environment with SARS-CoV-2 positive history were 4.23 (95% CI: 2.07, 8.63) times more likely to test positive for SARS-CoV-2 antibodies compared to those who did not know anyone with SARS-CoV-2 infection. Students who self-reported drinking alcohol more than one day a week were 1.79 (95% CI: 1.02, 3.16) more likely to have a positive antibody test compared to those who self-reported drinking alcohol equal to or less than one day a week. Similarly, students who socialized with more than four people when drinking alcohol were 2.21 (95% CI: 1.26, 3.90) times more likely to have a positive antibody test result compared to those who socialized with four or less people while drinking alcohol (Table 2). Because there were few to zero observations in many of the cells of race and outcome variables cross tabulations, we could not fit the unadjusted models with race as independent variable (Models with race variable did not converge).

#### 3.4.2 Subjective outcome: self-reported SARS-CoV-2 testing history

Similar results were observed for the associations between the aforementioned factors and the self-reported SARS-CoV-2 testing history outcome (Table 2). However, the point estimates for this outcome were measured more precisely with tighter confidence intervals, likely because of the larger sample size. For all associations, the magnitude of the prevalence ratios attenuated, yet they remained significantly and substantially above null. The largest attenuation in the magnitude occurred for knowing others who were infected variable, from 4.23 to 3.03 (95% CI: 2.04, 4.49).

### 3.5 Other analysis

We also evaluated the association between SARS-CoV-2 antibody laboratory test and self-reported SARS-CoV-2 testing history outcomes (Table 2). Of the 46 students who tested positive for SARS-CoV-2 antibodies and had complete self-reported testing data, 29 self-reported they had previously tested positive for an active SARS-CoV-2 infection, and 17 self-reported they had never tested positive for an active SARS-CoV-2 infection. The magnitude of the association was large [PR (95% CI): 16.73 (9.54, 29.33)]. Lastly, similar results were found in our sensitivity analysis of restricting the sample size to students below 30 years old (Appendix B).

## 4 Discussion

### 4.1 Key Results

In September 2020, near the beginning of the fall semester, seroprevalence of SARS-CoV-2 in our random study sample of Indiana University Bloomington undergraduate students was 4.6%, while the prevalence of students who self-reported SARS-CoV-2 infection history was 10.3%. We found that students who had Greek membership, had multiple partners, knew others in their immediate environment with SARS-CoV-2 infection, drank alcohol more than one day a week, and hanged out with more than four people when drinking were more likely to be tested positive for SARS-CoV-2 antibody test and self-report positive SARS-CoV-2 test history.

### 4.2 Interpretation

The SARS-CoV-2 seroprevalence among IUB undergraduate students was lower than the nationwide seroprevalence estimate in July 2020 (9.3%) (17) and higher than the Indiana statewide estimate in April 2020 (1.1%) (18). However, our findings are comparable to that of other large universities in the U.S. (2, 19). Selection bias might have influenced our seroprevalence estimate because we asked students not to attend their laboratory test appointment if they were tested positive for SARS-CoV-2 in the last two weeks, had been directed to isolate or quarantine, or were experiencing COVID-19 symptoms. This selection bias could have altered our seroprevalence estimate in either direction. However, because people with COVID-19 symptoms are more likely to have SARS-CoV-2 infection, the bias likely caused an underestimation of the true seroprevalence. Selection bias did not affect our estimate for the prevalence of students who self-reported ever having a SARS-CoV-2 positive test because this information was collected on the baseline online survey.

We also found that the prevalence of a self-reported SARS-CoV-2 positive test was higher than the seroprevalence collected via the laboratory test visit. At least some of this difference could be explained by the time lag between SARS-CoV-2 infection and antibody development and the fact that some infected individuals might never develop antibodies against the virus (20). However, it is less likely that the difference is because of immune memory loss in previously infected students. A recent study (yet to be peer reviewed) found that antibodies might last for years in recovered individuals (21, 22). This difference could also be due to the selection bias explained in the previous paragraph. Our team plans future analyses to further evaluate the reasons for the observed difference in the outcomes’ prevalence estimates.

Living in one of IU’s fraternities and sororities was a strong risk factor for seropositivity. Similarly, on other campuses, clusters of COVID-19 cases have been linked to Greek houses (23). Congregate living settings and the unofficial activities and gatherings (*e.g*., rush events) could possibly explain this strong association (23). We further found that students who were dating/hooking up with multiple people were more likely to self-report a positive SARS-CoV-2 test or have a positive SARS-CoV-2 antibody test result. To our knowledge, this is the first study that quantitatively evaluated this association. SARS-CoV-2 is primarily transmitted through direct contact with infected individuals or contaminated surfaces (*i.e*., fomite transmission) and/or exposure to large and small droplets that contain the virus (24), all of which are possible when students are dating/hooking up with multiple partners. Likewise, students who knew others with SARS-CoV-2 infection in their immediate environment were more likely to self-report a positive SARS-CoV-2 test or have a positive SARS-CoV-2 antibody test result. These students could also have been exposed to SARS-CoV-2 because of being in prolonged contact with the infected individuals.

Drinking alcohol more than once a week and drinking in groups of larger than four increased the likelihood of SARS-CoV-2 seropositivity. Young adults might adhere less strictly to COVID-19 prevention measures when drinking alcohol (8), probably because of cognitive distortion that follows drinking (25). In a social drinking event, students are likely to drink more when more friends are present (9), because of peer pressure, which can exacerbate the cognitive distortion and correspondingly cause further noncompliance with COVID-19 prevention measures. More importantly, presence of more friends in a drinking event brings in more possible sources of SARS-CoV-2 infections. Holding social events via online video-conferencing technologies, such as Zoom, or in settings where physical distancing is possible, avoiding excessive drinking, and drinking only with people who live in one’s household could help to reduce transmission of SARS-CoV-2 among college students.

### 4.3 Limitations and generalizability

In this study, because we used cross-sectional baseline data, we cannot assess temporal ordering between different study variables and outcomes. Even though confounding is usually a limitation in observational studies, adjusting for confounding was not necessary in the current study because our research questions were descriptive and predictive, and they were not about causal inference (26). Lastly, all data, except SARS-CoV-2 antibody laboratory test results, were collected through self-reported surveys. Different sources of bias, such as measurement and recall biases, could affect the quality of self-reported data. However, we found a very strong association between a positive SARS-CoV-2 antibody laboratory test result and a positive self-reported SARS-CoV-2 testing history, suggesting measurement bias may not be a significant concern for the self-reported data.

Despite the limitations, our study provides insight into the dynamics of SARS-CoV-2 seropositivity among college students and can help educational administrators and policy makers when developing future strategies for combating the pandemic in these settings. Particularly, as we used random sampling methods in this study to increase the external validity of our results, our findings may be applicable to other large universities in the U.S.

## Supporting information

STROBE

## Data Availability

Data are available upon request.

## Data Availability

Available upon request

## Funding

This study was supported by private contributions to the Indiana University Foundation. The United Arab Emirates provided the testing kits.

## Notes

**Conflict of interest:** None

### Competing Interest Statement

The authors have declared no competing interest.

### Clinical Trial

The current study design is cross-sectional. However, the parent study is a randomized controlled trial and is registered on ClinicalTrials.gov (ID: NCT04620798).

### Clinical Protocols

https://clinicaltrials.gov/ct2/show/NCT04620798

### Author Declarations

Indiana University's Human Subjects & Institutional Review Boards gave ethical approval for this study's protocol (Protocol #2008293852, Review Type: Expedited, Status: Approved).

