## Appendix A for "A cross-sectional analysis of demographic and behavioral risk factors of SARS-CoV-2 antibody positivity among a sample of U.S. college students"

| Table A1. Data codebook and question wording | | | |
| --- | --- | --- | --- |
| Variable name in the dataset | Question wording/variable description | Available options in the baseline survey/on REDCap instrument | Coding in the analysis |
| Outcomes |  |  |  |
| Objective Outcome |  |  |  |
| base_result_conf | Please enter the SARS-CoV-2 Antibody Test Results [for IgM and IgG] below.     - IgG Note: Positive if line is present - IgM Note: Positive if line is present | [Volunteers entered their readings for both IgG and IgM]  - Positive  - Negative | - If both IgG and IgM were negative, the antibody test result was negative  - If any of the IgG or IgM readings were positive, the antibody test result was positive |
| Subjective Outcome |  |  |  |
| ever_tested | Have you ever been tested for SARS-CoV-2 (COVID-19) before? Note: By this, we mean testing for active infections, usually done with a nasal swab or saliva test. | 0- No  1- Yes  98- Don't know^a^ |  |
| ever_positive | Have you ever tested positive for a SARS-CoV-2 (COVID-19) infection? | 0- No  1- Yes  98- Don't know^a^ |  |
| Self_report_positive_test | This variable was created during the data processing phase using ever_tested and ever_positive variables and shows if a participant has ever been tested positive for an active COVID-19 infection detection test. |  | - Yes [if ever_tested = Yes AND ever_positive = Yes]  - No [any condition other than above] |
| Predictors |  |  |  |
| age | What is your current age? | Text | - ≥22 years old  - <22 years old |
| born_sex | What sex were you assigned at birth, on your original birth certificate? | 1- Male  2- Female | 1- Male  2- Female |
| race | Which categories describe you? (Please choose all that apply) | [Checkbox]  1- White  2- Hispanic or Latinx or Spanish origin  3- Black or African-American  4- Asian  5- America Indian or Alaska Native  6- Middle Eastern or North African  7- Native Hawaiian or Other Pacific Islander  8- Some other race, ethnicity, or origin | - Asian  - Black  - Multi-racial  - Other  - White |
| race___2 | Hispanic or Latinx or Spanish origin | - Checked  - Unchecked | - Yes  - No |
| sch_year | What is your year in school? | 1- First year undergraduate  2- Second year undergraduate  3- Third year undergraduate  4- Fourth year undergraduate  5- Fifth year or more undergraduate | 1- First year undergraduate  2- Second year undergraduate  3- Third year undergraduate  4- Fourth and fifth year undergraduate |
| residence | Where do you currently live? | 1- On campus (in a dorm/residence hall)  2- Off campus apartment  3- Off campus house  4- At home with parents or other family members  5- Other | - On-campus  - Off-campus |
| greek_mmbr | What is your membership status with IUs fraternities and sororities? | 1- Member of an IFC fraternity  2- Member of a PHA sorority  3- Member of a MCGC fraternity  4- Member of a NPHC sorority  5- Member of another fraternity/sorority  6- Not a member of any fraternity/sorority | - Yes [if greek_mmbr equals any of the first five options]  - No [if greek_mmbr=6] |
| rel_status | Which of the following best describes your current relationship status? | 1- Single and not dating/hooking up with anyone  2- Single and dating/hooking up with multiple people  3- Single and dating/hooking up with one specific person  4- In a relationship but not living together  5- Living together but not married  6- Married and living together  7- Married but not living together | - Multiple partners [if rel_status=2]  - Single partner [if rel_status equals 3, 4, 5, or 6]  - No partners [if rel_status = 1] |
| symptoms | [Showed the question ONLY if:  ever_positive = 1]  If yes, how would you describe the symptoms of your infection? | 1- Asymptomatic  2- Mild  3- Moderate  4- Severe  98- Don't know^a^ | - Yes [if symptoms equals 2, 3, or 4]  - No [if symptoms = 1] |
| know_others_infected | Do you know people in your immediate social environment who are or have been infected with COVID -19 (suspected or confirmed)? | 0- No  1- Yes  98- Don't know^a^ | 0- No  1- Yes |
| alc_freq_v2 | How many days a week do you usually drink alcohol? | 0- 0 days or don't drink alcohol  1- 1 day  2- 2 days  3- 3 days  4- 4 days  5- 5 days  6- 6 days  7- 7 days | - Equal or less than 1 [if alc_freq_v2<=1]  - More than 1 [alc_freq_v2>1] |
| drink_people_n | On a typical night, how many people do you usually hang out with when you are drinking alcohol? | text (number, Min: 0, Max: 1000) | - Equal or less than 4 [if drink_people_n<=4]  - More than 4 [drink_people_n>4] |
| a. All the Don’t know responses were coded as missing | | | |
