## Appendix B for "A cross-sectional analysis of demographic and behavioral risk factors of SARS-CoV-2 antibody positivity among a sample of U.S. college students"

| **Table B1. Bivariate prevalence ratios for the associations between risk factors and positive** **SARS-CoV-2 antibody test and self-reported history of positive SARS-CoV-2 test, restricted to participants below 30 years old** | | |
| --- | --- | --- |
| **Predictor** | **Outcomes** | |
|  | **Positive SARS-CoV-2 Antibody Test**  **(objective outcome)** | **Self-reported History of Positive SARS-CoV-2 Test (subjective outcome)** |
|  | PR (95% CI) | PR (95% CI) |
| **Age** | | |
| ≥22 AND <30 years old | 0.75 (0.24, 2.37) | 0.68 (0.34, 1.36) |
| <22 years old | Ref. | Ref. |
| **Sex at birth** | | |
| Female | 1.44 (0.77, 2.69) | 0.88 (0.63, 1.23) |
| Male | Ref. | Ref. |
| **Race** | | |
| White | 0.89 (0.46, 1.72) | 1.33 (0.87, 2.05) |
| Other | Ref. | Ref. |
| **Hispanic or Latinx Ethnicity** | | |
| Yes | 1.84 (0.81, 4.19) | 1.05 (0.57, 1.93) |
| No | Ref. | Ref. |
| **Race (reclassified)** | | |
| Hispanic | 1.82 (0.79, 4.16) | 0.98 (0.53, 1.80) |
| Non-white non-Hispanic | 0.89 (0.32, 2.44) | **0.41 (0.19, 0.92)** |
| White non-Hispanic | Ref. | Ref. |
| **Year in school** | | |
| 1^st^ | 1.02 (0.49, 2.11) | 1.09 (0.69, 1.70) |
| 2^nd^ | 0.89 (0.42, 1.89) | 1.13 (0.73, 1.77) |
| 3^rd^ | 0.69 (0.31, 1.54) | 0.86 (0.53, 1.38) |
| 4^th^ or 5^th^ | Ref. | Ref. |
| **Residence** | | |
| On -campus | 1.10 (0.61, 1.97) | 1.27 (0.91, 1.78) |
| Off-campus | Ref. | Ref. |
| **Greek membership** | | |
| Yes | **3.40 (1.97, 5.89)** | **2.42 (1.75, 3.34)** |
| No | Ref. | Ref. |
| **Relationship Status** | | |
| Multiple partners | **2.51 (1.19, 5.32)** | **2.18 (1.43, 3.32)** |
| Single partner | 1.00 (0.53, 1.87) | 0.80 (0.55, 1.16) |
| No partner | Ref. | Ref. |
| **Self-report positive test (Subjective outcome)** | | |
| Yes | **17.63 (9.93, 31.29)** | -- |
| No | Ref. | -- |
| **Symptomatic ^a^** | | |
| Yes | 1.51 (0.65, 3.51) | -- |
| No | Ref. | -- |
| **Know others who were infected** | | |
| Yes | **4.72 (2.23, 9.99)** | **3.01 (2.03, 4.46)** |
| No | Ref. | Ref. |
| **Number of days/week drinking alcohol** | | |
| More than 1 day | **1.73 (0.98, 3.05)** | **1.60 (1.14, 2.24)** |
| 1 day or less | Ref. | Ref. |
| **Number of people hanging out with while drinking** | | |
| More than 4 people | **2.31 (1.30, 4.10)** | **1.77 (1.26, 2.48)** |
| 4 people or less | Ref. | Ref. |
| a. Displayed if self-report positive test = Yes  Boldface indicates *p*<0.05 | | |
